# Regional Variation after Cardiac Surgery in Adults with Congenital Heart Disease in the US

**DOI:** 10.64898/2025.12.17.25342525

**Authors:** Carlos E. Diaz-Castrillon, Victor Morell, Kenneth J. Smith, Melita Viegas, Mario Castro-Medina, Ibrahim Sultan, Arvind K. Hoskoppal, Yisi Wang, Jennifer Nelson

**Author notes:** Correspondence and Reprint Requests: Carlos E. Diaz-Castrillon; Department of Cardiothoracic Surgery; UPMC Children’s Hospital of Pittsburgh, 4401 Penn Avenue, Central Plant Building, Pittsburgh, PA 15224. Funding: none. Disclosures: none.

## Abstract

**Introduction:** We describe regional variation in perioperative care among adults with congenital heart disease (ACHD) undergoing cardiac surgery in the United States.

**Methods:** Analysis using the 2016–2020 Nationwide Inpatient Sample, including admissions of ≥18 years old patients with congenital heart ICD-10 diagnosis codes and cardiac surgical procedure codes. Outcomes included hospitalization costs, length of stay, high-resource utilization (HRU), and in-hospital mortality. Adjusted multilevel regressions estimated observed-to-expected (O/E) ratios and risk-standardized (RS) outcomes.

**Results:** Among 83,800 admissions, the median age was 59 years (IQR 48-67) and the most common diagnosis was congenital aortic insufficiency (AI) with 58.7% (n=49,149). Elective admissions represented 76.5% (n=64,115) admissions; valve replacements were coded in 68.1% (n=57,065) admissions, followed by CABG (22.0%, n=18,420). Regional distribution was 21% West, 20% Northeast, 27% Midwest, and 31% South. Midwest had a higher proportion of valve repairs whereas Northeast had a higher valve replacement procedure. RS-HRU rate raged from 14.8% (95% CI 13.0–16.7) in the Midwest to 20.4% (18.5–22.3) in the South. RS-costs varied from $49,303 (95% CI 47,944–50,665) in the Northeast to $59,899 (95% CI 58,432–61,355) in the West. In-hospital mortality did not significantly differ by region. Sensitivity analysis excluding congenital AI admissions showed increased resource utilization and mortality.

**Conclusion:** Substantial regional variation exists in ACHD resource utilization. Future studies will focus on specific procedure types and hospital characteristics to uncover potential region-specific strategies to optimize ACHD care as healthcare moves toward value-based models aligned with regional demands.

## Introduction

Advances in perioperative care for pediatric and congenital heart disease (CHD) have significantly improved survival, leading to a growing adult congenital heart disease (ACHD) population. Between 1990 and 2019, the CHD population increased by 28%, with notable growth among adults aged 15–49 (42%) and 50–69 (117%). (1) This expansion has heightened demand for resource-intensive care, including surgical interventions (2)

Compared to the non-ACHD population, ACHD patients face higher resource utilization, requiring more frequent emergency care, prolonged postoperative management, specialized rehabilitation, and lifelong monitoring. (3–6) Furthermore, prior studies have shown that ACHD patients undergoing coronary artery bypass grafting (CABG) not only incur higher hospital costs but also experience worse outcomes compared to patients without CHD undergoing the same procedure. (7)

Despite this, comprehensive data on regional differences in resource utilization and costs after ACHD cardiac surgery in the U.S. are lacking. Describing the interplay between each region’s case-mix and related demands on cardiac surgical services across regions could provide insights to healthcare policymakers or healthcare leaders in allocating and managing resources. Thus, the purpose of this study was to: 1) describe regional differences in resource utilization and hospitalization costs and, 2) determine the burden of cardiac surgical ACHD care in each region using risk-adjustment to account for differences in regional case-mix.

## Methods

This study was conducted using the 2016-2020 Nationwide Inpatient Sample (NIS) database, the largest publicly available all-payer inpatient healthcare database designed to produce U.S. regional and national estimates of inpatient utilization, access, cost, quality, and outcomes.(8) The NIS is part of a family of databases and software tools developed for the Healthcare Cost and Utilization Project (HCUP). NIS approximates a 20-percent stratified sample of all discharges from U.S. community hospitals, excluding rehabilitation and long-term acute care hospitals, including 48 states plus the District of Columbia. This study was deemed exempt from further review by the University of Pittsburgh Institutional Review Board.

### Study Population and baseline characteristics

We analyzed hospital admissions of adults (≥18 years) with CHD using ICD-10 codes from Q20–Q26, covering congenital malformations of the circulatory system. Admissions involving surgical procedures of the heart and great vessels were identified, excluding specific diagnostic codes for rheumatic valve disease, bicuspid valves, and hospitals with fewer than one case per year. (**Supplementary Figure 1**) Baseline characteristics included age, sex, Elixhauser Comorbidity Index, race/ethnicity, expected payer, and median household income, as well as hospital characteristics such as teaching status, size, and volume.

### Outcomes

The primary outcomes were in-hospital mortality and hospitalization costs, adjusted for inflation to 2023 US dollars and regional labor expenses using the area-specific wage index. Admissions with costs at the lowest 1% were excluded to enhance reliability. Secondary outcomes included high-resource utilization (HRU), defined as the presence of one or more intensive interventions (e.g., defined by the presence of one or more of the following criteria: mechanical ventilation > 24 hours, hemodialysis, tracheostomy, cardiopulmonary resuscitation, blood transfusion, ECMO or temporary mechanical circulatory support.), and length of stay (LOS).

### Analytical Approaches

Multivariable multilevel regression models were fitted to quantify the association of the region with the outcomes of interest. Adjustments were made for patient-level characteristics (age, sex, comorbidities, race/ethnicity, payer type, diagnosis, median household income), hospital-level factors (teaching status, size, volume, ownership), and procedural variables (procedure type, number of codes, complexity). The Risk Adjustment for Congenital Heart Surgery-2 (RACHS-2) was included as a complexity indicator. (9) The RACHS-2 for ICD-10(c) is an empirically derived, risk adjustment model that has been validated in two separate administrative data sources and compared to locally held clinical registry data.

To evaluate regional variation in outcomes, we used two complementary approaches. First, we estimate predicted outcomes using the “*margins atmeans”* command, which reflect region-specific effects while holding covariates at their mean values. These adjusted predictions allow for comparisons assuming an identical case mix across regions and help isolate differences attributable to regional practice or system-level factors. A second set of estimtates were predicted using the “*margins asobserved*” command, which account for each patient’s actual risk profile. These represent real-world risk-adjusted outcomes, reflecting the actual case mix within each region.

These models were also used to calculate observed-to-expected (O/E) ratios for each region. This metric allows for fair comparisons by highlighting differences in performance relative to expected outcomes based on case mix.

National estimates were calculated using NIS discharge weights. Descriptive statistics summarized patient and hospital characteristics. Continuous variables were reported as medians with interquartile ranges (IQR), while categorical variables were presented as proportions. Group comparisons employed chi-square tests for categorical and Kruskal-Wallis or ANOVA for continuous variables, based on distribution. For hospitalization costs and length of stay, we used multilevel generalized linear models (GLM) with Gamma and Poisson distributions, respectively, and multilevel logistic regression for in-hospital mortality and high-resource utilization (HRU). We used Stata® 18 (StataCorp LLC, College Station, TX).

## Results

A total of 83,800 hospital admissions were identified. The median age was 59 years (IQR 48-67), and 30.7% (n=25,705) were women. Regionally, 20.3% (n=16,985) of admissions occurred in the Northeast, 21.0% (n=17,625) in the West, 27.2% (n=22,755) in the Midwest, and 31.5% (n=26,435) in the South. (**Table 1, supplementary table 1**). Median hospital volume was 11 (IQR 5-23) admissions/year. (**Figure 1**). Private insurance covered 51.1% (n=42,780) of admissions, while 43.4% (n=36,330) were covered by public insurance, with highest proportion of private insurance coverage in the Midwest with 53.6% of admissions. Among all regions, private not-for-profit hospitals accounted for most admissions (84.3%), with the highest proportion in the Midwest (92.7%) and the lowest in the South (72.3%). Private investor-owned hospitals were least common (6.6%), with higher concentrations in the South (13.1%) and West (6.2%). (**Supplementary table 1** and **figure 2**)

**Figure 1.**
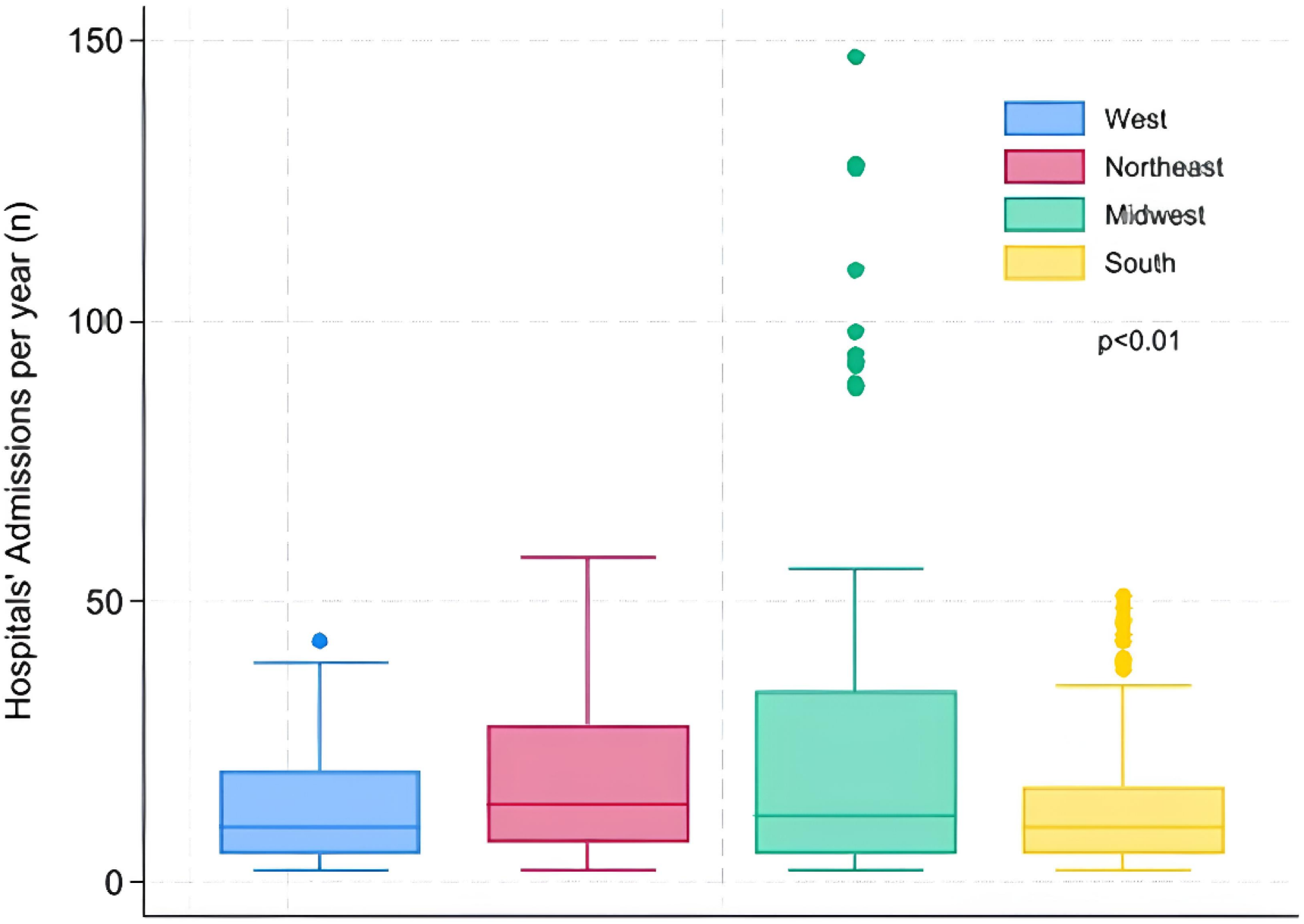
Annual hospital admission volume per year. The West has a median hospital admission volume of 10 cases per year (IQR 5–20), the Northeast 14 (IQR 7–28), the Midwest 12 (IQR 5–34), and the South 12 (IQR 5–34).

**Figure 2.**
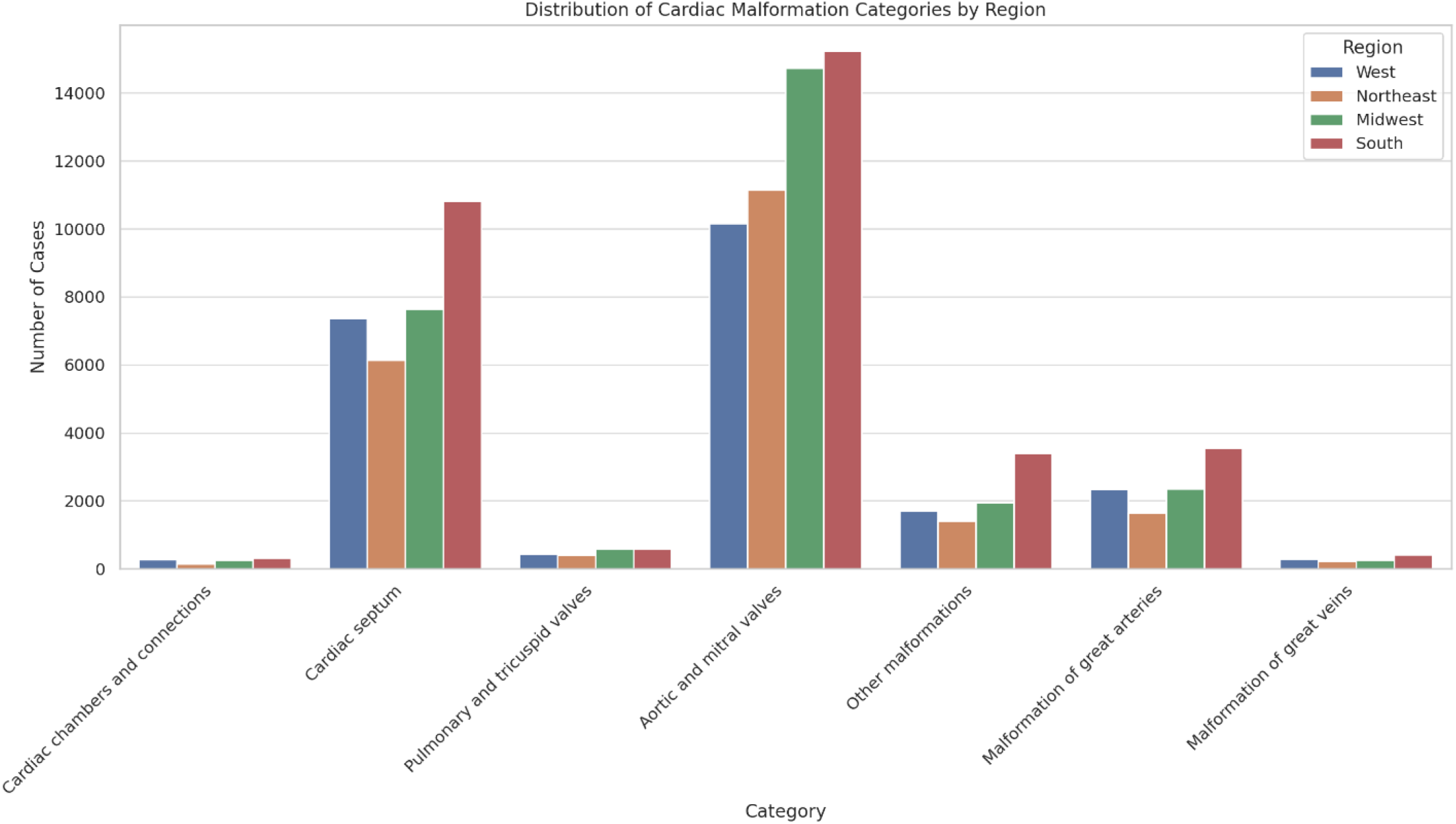
Distribution of congenital cardiac malformations ICD-10 diagnosis categories by region.

**Figure 3.**
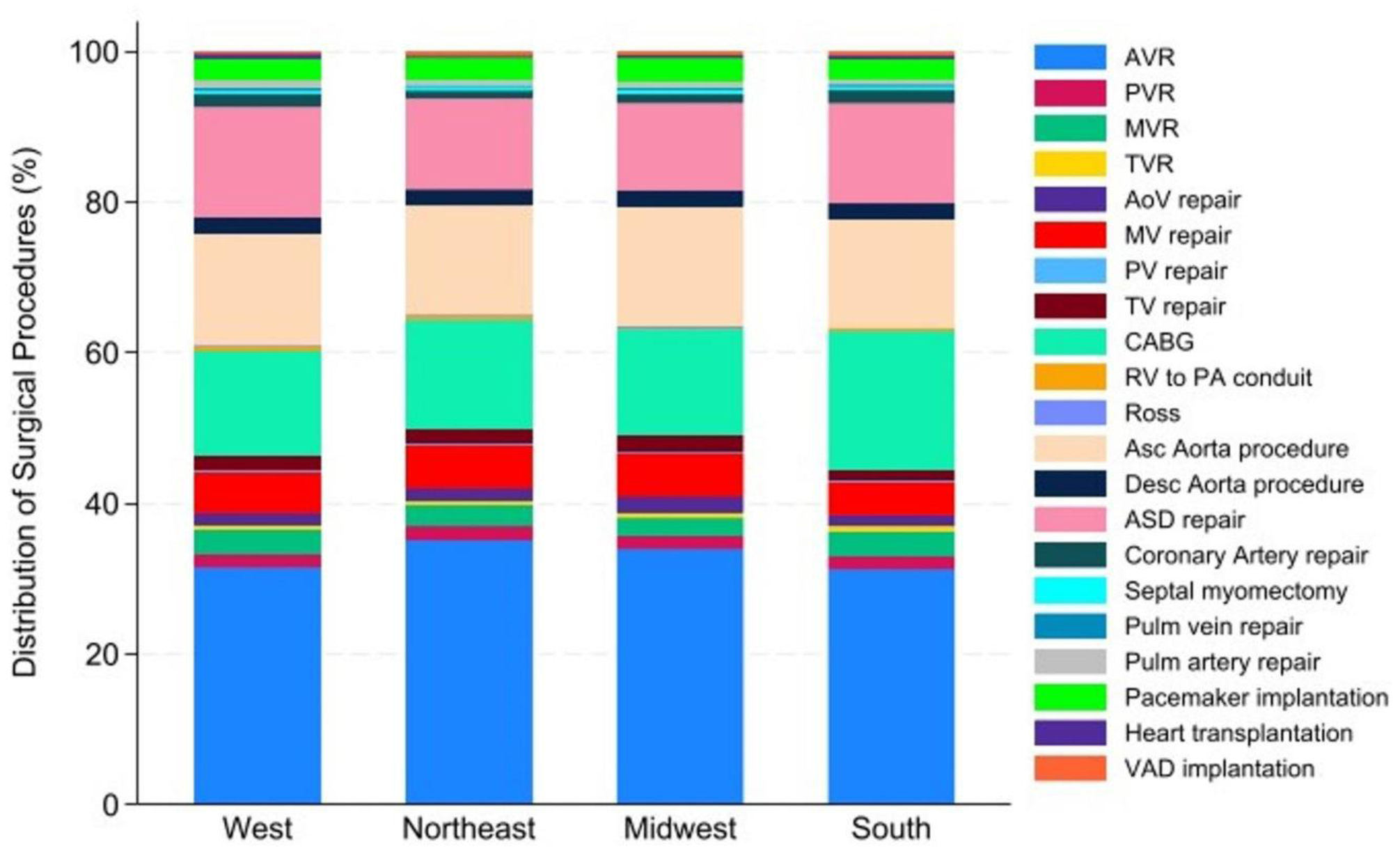
Regional Distribution of specific cardiac procedures.

**Table 1.** ACHD Cardiac Surgery Outcomes and Resource Utilization.

| Region | West<br>N= 17,625 | Northeast<br>N= 16,985 | Midwest<br>N= 22,755 | South<br>N= 26,435 | Total<br>N=83,800 | p<br>value |
| --- | --- | --- | --- | --- | --- | --- |
| **Lenth of stay, median (IQR)** | 6 days (5-9) | 6 days (5-10) | 6 days (5-9) | 7 days (5-10) | 6 days (5-9) | 0.001 |
| **Percentile 90<sup>th</sup> LOS, n (%)** | 1,880 (9.1%) | 2,070 (10.3%) | 2,085 (7.9%) | 3,800 (11.8%) | 9,090 (10.8%) | 0.001 |
| **Mortality, n (%)** | 245 (1.4%) | 195 (1.1%) | 290 (1.3%) | 485 (1.8%) | 1,215 (1.4%) | 0.07 |
| **Median hospitalization costs (IQR)** | \$54,943<br>(\$42,945-<br>\$72,282) | \$44,152<br>(\$34,404-<br>\$60,598) | \$49,509<br>(\$38,236-<br>\$66,374) | \$55,065<br>(\$43,269-<br>\$74,564) | \$51,421<br>(\$39,908 -<br>\$69,186) | 0.001 |
| **Percentile 90<sup>th</sup> Costs, n (%)** | 1,910 (10.8%) | 1,280 (7.5%) | 1,950 (8.6%) | 3,305 (12.5%) | 8,445 (10.1%) | 0.001 |
| **HRU, n (%)** |  |  |  |  |  |  |
| **Mech ventilation 24-96hrs** | 355 (2.0%) | 350 (2.1%) | 530 (2.3%) | 805 (3.0%) | 2,040 (2.4%) | 0.01 |
| **Mech ventilation>96hrs** | 340 (1.9%) | 255 (1.5%) | 360 (1.6%) | 550 (2.1%) | 1,505 (1.8%) | 0.72 |
| **Tracheostomy** | 70 (0.4%) | 105 (0.6%) | 85 (0.4%) | 135 (0.5%) | 395 (0.5%) | 0.34 |
| **CPR** | 115 (0.7%) | 75 (0.4%) | 225 (1.0%) | 145 (0.5%) | 560 (0.7%) | 0.04 |
| **Renal Replacement Therapy** | 170 (1.0%) | 110 (0.6%) | 200 (0.9%) | 235 (0.9%) | 715 (0.9%) | 0.31 |
| **Mechanical circulatory support** | 570 (2.7%) | 535 (2.7%) | 700 (2.7%) | 1,180 (3.7%) |  | 0.01 |
| **ECMO** | 55 (0.3%) | 25 (0.1%) | 70 (0.3%) | 105 (0.4%) | 255 (0.3%) | 0.40 |
| **Blood Transfusion** | 2,845 (16.1%) | 2,690 (15.8%) | 2,545 (11.2%) | 5,035 (19.0%) | 13,115 (15.7%) | 0.001 |
| **Any HRU, n (%)** | 3,780 (21.4%) | 3,540 (20.8%) | 3,775 (16.6%) | 6,695 (25.3%) | 17,790 (21.2%) | 0.001 |
| **>1 HRU procedure, n (%)** | 2,875 (16.3%) | 2,570 (15.1%) | 3,260 (14.3%) | 4,235 (16.0%) | 12,940 (15.4%) | 0.02 |
| **Total count of HRU, n (%)** |  |  |  |  |  | 0.001 |
| **0** | 13,845 (78.6%) | 13,445 (79.2%) | 18,980 (83.4%) | 19,740 (74.7%) | 66,010 (78.8%) |  |
| **1** | 3,290 (18.7%) | 3,165 (18.6%) | 3,150 (13.8%) | 5,670 (21.4%) | 15,275 (18.2%) |  |
| **2** | 355 (2.0%) | 285 (1.7%) | 410 (1.8%) | 825 (3.1%) | 1,875 (2.2%) |  |
| **3 or more** | 135 (0.8%) | 90 (0.5%) | 215 (0.9%) | 200 (0.8%) | 640 (0.8%) |  |

### ICD-10 Congenital Malformation Categories

Diagnosis codes from the Q23 category (aortic and mitral valve malformations) were the most frequently observed across the cohort, accounting for 60.3% of all admissions (n = 50,555), with the lowest regional prevalence in the South (56.9%) (**Figure 2 and supplementary table 1-2**). Notably, congenital AI overwhelmingly predominated within this category, representing 97.3% of Q23 diagnoses (n = 49,195). The second most common diagnostic group was Q21 (cardiac septal defects), present in 22.3% of admissions (n = 18,655), with the highest prevalence in the West (26.0%) and the lowest in the Midwest (19.7%) (p < 0.001). Among these, ASD comprised the majority, representing 83% of Q21-coded admissions.

Pulmonary congenital anomalies, including pulmonary atresia, stenosis, insufficiency, and other valve malformations, were recorded in 56.4% of Q22 admissions (n = 1,045), with relatively uniform distribution across regions (West 60.0%, Northeast 64.0%, Midwest 43.6%, South 61.6%; p = 0.07). Tetralogy of Fallot accounted for 4.1% (n = 760) of the Q21 category, without differences between regions. Conotruncal anomalies—including truncus arteriosus, DORV, and TGA—collectively accounted for approximately 630 cases (66.3% of Q20). Coronary artery anomalies represented 70.0% of Q24 diagnoses and 7.3% of the total cohort (n = 6,155), with the highest frequency in the South (77.9%) and the lowest in the Northeast (63.3%) (p = 0.002).

### Regional Variation in Surgical procedures

Elective admissions accounted for 76.5% (n = 64,115) of all admissions, with the highest proportion in the West (75.8%) and lowest in the Northeast (74.4%) (p < 0.001) (**Supplementary table 3**). Valve replacement was the most frequently performed procedure, observed in 68.1% (n = 57,065) of all admissions. By region, the proportion of admissions involving valve replacement was highest in the Northeast (71.8%, n = 12,195) and lowest in the South (65.5%, n = 17,315) (p < 0.001). Aortic valve replacement (AVR) alone accounted for 33.2% (n = 19,445) of admissions, ranging from 36.2% in the Northeast to 32.2% in the South (p = 0.26).

CABG was performed in 22.0% (n = 18,420) of admissions, with the highest frequency in the South (25.3%, n = 6,680) and lowest in the Northeast (20.4%, n = 3,465) (p < 0.001). Valve repair procedures were performed in 13.8% (n = 11,550) of admissions. Regional distribution showed the highest rate in the Midwest (15.5%) and the lowest in the South (11.9%) (p < 0.001).

### Outcomes and Resource Utilization

#### Unadjusted Outcomes

In-hospital mortality was 1.1% in the Northeast, 1.3% in the Midwest, 1.4% in the West, and 1.8% in the South (p=0.06) (**Table 1**). HRU procedures rate occurred in 21.2% (n=17,790) of admissions, with the highest frequency in the South (24.2%) and the lowest in the Midwest (18.8%). The median LOS for the full cohort was 6 days (IQR 5–10), with no significant regional differences in unadjusted medians. The median unadjusted hospitalization cost was $51,421 (IQR $39,908–$69,186), highest in the West ($55,245) and lowest in the Northeast ($46,387).

#### Adjusted Outcomes

After adjustment for patient demographics, comorbidities, and procedural complexity, there were no statistically significant regional differences in in-hospital mortality (**Table 2**). However, modest variation was observed between estimates predicted at means (standardized covariates across regions) versus those predicted as observed (based on each region’s actual case mix), suggesting that regional differences in patient profiles may contribute to higher mortality estimates observed across the regions (**Table 3**).

**Table 2.** Adjusted analysis of mortality, costs, length of stay (LOS), and admissions with high-resource utilization (HRU) across different U.S. regions relative to the West (reference region). The cost and LOS are calculated as percentage changes. Mortality and HRU are odds ratios.

| **Metric** | **Northeast** | **Midwest** | **South** |
| --- | --- | --- | --- |
| **Mortality** | 1.06 (0.67 – 1.68) | 1.09 (0.70 – 1.69) | 1.14 (0.74 – 1.74) |
| **Costs** | –17.7% (95% CI: –20.3% to –14.9%) | –3.1% (95% CI: –5.8% to –0.2%) | –0.4% (95% CI: –3.1% to +2.3%) |
| **LOS** | +6.8% (95% CI: +2.7% to +11.1%) | –0.9% (95% CI: –4.4% to +2.8%) | +6.4% (95% CI: +2.5% to +10.4%) |
| **HRU** | 1.14 (0.91 to 1.41) | 0.81 (0.63 to 1.02) | 1.22 (1.03 to 1.53) |
| **Without admissions with Congenital Aortic insufficiency ICD-10 code** |  |  |  |
| **Mortality** | 1.31 (0.77 – 2.24) | 1.06 (0.62 – 1.78) | 1.34 (0.85 – 2.10) |
| **Costs** | –16.8% (95% CI: –20.5% to –13.0%) | –3.8% (95% CI: –7.6% to +0.3%) | –0.7% (95% CI: –4.3% to +3.0%) |
| **LOS** | +10.2% (95% CI: +3.7% to +17.1%) | –0.9% (95% CI: –6.5% to +5.0%) | +7.5% (95% CI: +1.7% to +13.5%) |
| **HRU** | 1.13 (0.85 to 1.52) | 0.82 (0.63 to 1.08) | 1.22 (0.95 to 1.58) |

**Table 3:** Risk-standardized outcomes among ACHD during cardiac surgical admissions. Estimations derived from multilevel regression analysis.

| **Outcome** | **Region** | **Predicted at means-Based on**<br>**constant covariates across regions** | **Predicted as observed-Based on**<br>**each region's actual case mix** |
| --- | --- | --- | --- |
| **Cost (USD)** | **Northeast** | \$49,303 (47,944–50,665) | \$73,039 (64,256–81,825) |
| | **Midwest** | \$58,070 (56,666–59,473) | \$86,025 (75,597–96,453) |
| | **South** | \$59,642 (58,363–60,951) | \$88,354 (77,746–98,963) |
| | **West** | \$59,899 (58,432–61,355) | \$88,736 (78,026–99,445) |
| **Length of Stay (days)** | **Northeast** | 7.9 (7.7–8.2) | 8.8 (8.5–9.1) |
|  | **Midwest** | 7.4 (7.2–7.6) | 8.2 (7.9–8.4) |
|  | **South** | 7.9 (7.7–8.2) | 8.8 (8.5–9.1) |
|  | **West** | 7.5 (7.2–7.7) | 8.3 (8.0–8.5) |
| **In-Hospital Mortality** | **Northeast** | 0.5% (0.2–0.6%) | 1.3% (0.9–1.7%) |
|  | **Midwest** | 0.4% (0.3–0.6%) | 1.4% (1.0–1.8%) |
|  | **South** | 0.5% (0.3–0.6%) | 1.5% (1.1–1.8%) |
|  | **West** | 0.4% (0.2–0.6%) | 1.3% (0.9–1.7%) |
| **HRU Rate** | **Northeast** | 19.4% (17.3–21.5%) | 20.6% (18.6–20.5%) |
|  | **Midwest** | 14.8% (13.0–16.7%) | 16.1% (14.3–17.9%) |
|  | **South** | 20.4% (18.5–22.3%) | 21.6% (19.9–23.4%) |
|  | **West** | 17.3% (15.2–19.4%) | 18.6% (16.6–20.5%) |

**Table 4.** Adjusted estimations for specific procedures excluding ACHD admissions with congenital aortic insufficiency and isolated ASD as diagnosis.

|  | West | Northeast | Midwest | South |
| --- | --- | --- | --- | --- |
| ***Mortality*** |  |  |  |  |
| **ASD repair** | 1.1% (0.6 – 1.6) | 1.4% (0.7 – 2.0%) | 1.2% (0.6 – 1.7) | 1.5% (0.9 – 2.0) |
| **Valve repair** | 1.8% (0.9 - 2.6) | 2.2% (1.1 - 3.3) | 1.9% (0.9 - 2.8) | 2.3% (1.3 - 3.3) |
| **Valve replacement** | 1.8% (1.0 - 2.6) | 2.3% (1.2 - 3.2) | 2.0% (1.1 - 2.8) | 2.4% (1.6 – 3.2) |
| **CABG** | 2.1% (1.2 - 3.0) | 2.6% (1.4 - 3.8) | 2.2% (1.2 - 3.2) | 2.7% (1.8 - 3.7) |
| **Heart transplant** | 5.1% (0.6 - 9.6) | 6.2% (0.7 - 11.6) | 5.4% (0.5 - 10.2) | 6.5% (1.0 - 12.0) |
| ***HRU*** |  |  |  |  |
| **ASD repair** | 19.5% (16.2 – 22.7) | 21.2% (17.6 – 24.8) | 17.1% (14.1 – 19.9) | 22.1% (19.3 – 25.3) |
| **Valve repair** | 21.7% (18.6 - 25.0) | 23.6% (19.9 - 27.4) | 23.6% (16.1 - 27.4) | 24.9% (21.7 - 28.1) |
| **Valve replacement** | 22.8% (19.6 - 25.9) | 24.7% (21.2 - 28.3) | 20.1% (17.4 - 23.1) | 26.1% (23.2 - 28.9) |
| **CABG** | 22.8% (19.3 - 26.3) | 24.7% (20.9 - 28.5) | 20.1% (16.9 - 23.2) | 26.1% (22.9 - 29.1) |
| **Heart transplant** | 46.8% (37.9 - 55.7) | 50.3% (41.2 - 59.4) | 44.6% (35.7 - 53.4) | 50.5% (41.8 - 59.2) |
| ***LOS*** |  |  |  |  |
| **ASD repair** | 7.9 days (6.6- 9.1) | 10.9 days (9.1 – 12.8) | 8.6 (7.2 – 10.1) | 9.6 days (8.5 – 10.7) |
| **Valve repair** | 9.4 days (8.4 - 9.9) | 10.7 days (10.1 - 11.3) | 8.9 days (8.5 - 9.3) | 10.0 days (9.6 - 10.4) |
| **Valve replacement** | 9.9 days (9.4 - 10.3) | 11.2 days (10.6 - 11.8) | 9.3 days (8.8 - 9.7) | 10.4 days (10.0 - 10.8) |
| **CABG** | 10.1 days (9.6 - 10.6) | 11.6 days (10.9 - 12.3) | 9.5 days (9.0 – 10.0) | 10.7 days (10.3 - 11.2) |
| **Heart transplant** | 37.7 days (36 - 39) | 40.2 days (38.2 - 42.1) | 36.7 days (35.4 - 38.2) | 41.2 days (39.5 - 4.2) |
| ***Costs*** |  |  |  |  |
| **ASD repair** | \$63,119 (56,639 – 69,599) | \$55,830 (49,542 - 62,119) | \$60,693 (54,327 – 67,059) | \$59,550 (55,052 – 64,048) |
| **Valve repair** | \$68,431 (65,698 - 71,163) | \$60,680 (58,068 - 63,304) | \$65,925 (63,347 - 68,503) | \$70,368 (67,942 - 72,795) |
| **Valve replacement** | \$73,500 (70,734 - 76,265) | \$65,175 (62,500 - 67,849) | \$70,809 (68,213 - 73,404) | \$75,581 (73,231 - 77,931) |
| **CABG** | \$68,655 (66,001 - 71,309) | \$60,879 (58,320 - 63,438) | \$66,141 (63,662 - 68,621) | \$70,599 (68,352 - 72,846) |
| **Heart transplant** | \$190,260 (173,474 - 207,046) | \$168,711 (153,530 - 183,891) | \$183,294 (167,214 - 199,375) | \$195,648 (178,901 - 212,395) |

In contrast, significant regional variation was identified in resource utilization and hospitalization costs. Compared to the West, the South had higher odds (OR: 1.22; 95% CI 1.03 to 1.53). Regional differences in LOS were modest; LOS was slightly shorter in the Midwest (–8.7%; 95% CI: –13.4% to –3.9%) and the Northeast (–5.6%; 95% CI: –10.6% to –0.5%) compared to the West (Table 2).

On average, Northeast hospitalization costs were significantly lower in the Northeast (–17.7%; 95% CI: –20.3% to –14.9%) and modestly lower in the Midwest (–3.1%; 95% CI: –5.8% to –0.2%) relative to the West (**Table 2**). When evaluated in absolute terms, estimates based on observed case mix revealed that the Northeast remained the least costly region ($73,039; 95% CI: $64,256–81,825), while the West remained the most expensive ($88,736; 95% CI: $78,026–99,445) (**Table 3**). Again, predicted estimations based on each region’s actual case mix were overall higher, suggesting that patient profiles are associated with higher resource utilization.

These patterns were supported by the observed-to-expected (O/E) analysis (**Supplementary Table 4**). Both the Midwest (O/E: 1.10; 95% CI: 1.02–1.17) and South (O/E: 1.08; 95% CI: 1.02–1.13) had higher-than-expected HRU rates after adjusting for case mix. LOS differences were minimal across regions, with O/E ratios near 1.0, indicating comparable lengths of stay. Although all regions had cost O/E ratios below 1.0, indicating lower-than-expected hospitalization costs, the Northeast exhibited the most precise estimate, with a narrow confidence interval (O/E: 0.86, 95% CI: 0.74–0.99), suggesting more consistent performance relative to expected values.

### Sensitivity analysis

After excluding congenital AI admissions, subgroup comprised 48,600 admissions with a median age of 58 years (IQR 44–67), distributed regionally as follows: 22% West, 18% Northeast, 24% Midwest, and 35% South. In-hospital mortality estimates increased across all regions suggesting higher complexity, but overlapping confidence intervals continued to suggest no statistically significant differences. Regional patterns in adjusted outcomes remained consistent in resource utilization (**Table 2 and supplementary table 5**). The Northeast consistently demonstrated the lowest adjusted cost estimates.

### Procedure-Specific Outcomes excluding Congenital AI and isolated ASD

ASD repair was associated with the lowest mortality (1.1%–1.5%) and costs, though the Northeast exhibited a significantly longer length of stay (LOS: 10.9 days [95% CI: 9.1–12.8]) compared to the West (7.9 days [6.6–9.1]). For valve repair (mortality 1.8%–2.3%), costs were significantly lower in the Northeast ($60,600 [95% CI: $58,068–$63,304]) and highest in the South ($70,368 [95% CI: $67,942–$72,795]). In valve replacement admissions, while mortality ranged from 2.0% in the Midwest to 2.6% in the Northeast without statistically significant differences, the Northeast had significantly longer LOS (11.2 days [95% CI: 10.6–11.8]) and lower costs ($65,175 [95% CI: $62,500–$67,849]), whereas the South had significantly higher costs ($75,581 [95% CI: $73,231–$77,931]) and higher HRU (26.1%), with minimal overlap compared to the Midwest (20.1%).

CABG admissions showed similar trends: mortality ranged from 2.1% to 2.7% without significant regional differences, but LOS was significantly longer in the Northeast (11.6 days [95% CI: 10.9–12.3]) than in the Midwest (9.5 days [95% CI: 9.0–10.0]), and costs were significantly lower in the Northeast ($60,879 [95% CI: $58,320–$63,438]) compared to the South ($70,599 [95% CI: $68,352–$72,846]). Heart transplantation had the highest resource utilization and mortality (5.1%–6.5%). While mortality differences were not statistically significant, LOS was significantly longer in the South (41.2 days [95% CI: 39.5–42.9]) compared to the Midwest (36.7 days [95% CI: 35.4–38.2]), though cost differences across regions were not statistically significant.

## Comment

This national analysis provides a comprehensive assessment of regional variation in outcomes and resource utilization among ACHD undergoing cardiac surgery in the United States. The principal finding is that, despite substantial regional differences in case mix and procedural patterns, risk-adjusted in-hospital mortality did not differ significantly across regions, whereas resource utilization—particularly hospitalization costs and high-resource utilization (HRU)—varied meaningfully. These findings suggest that regional differences in efficiency and care delivery models, rather than survival outcomes, represent the dominant source of variation in ACHD surgical care.

Procedural patterns differed across regions, likely reflecting variation in referral pathways, institutional expertise, and diagnostic composition. The Midwest demonstrated a higher proportion of elective admissions and valve repair procedures, while the Northeast had greater use of valve replacement, and the South showed higher representation of CABG. In contrast, the distribution of major congenital diagnostic categories—such as pulmonary valve anomalies, tetralogy of Fallot, and conotruncal defects—was largely similar across regions, indicating that differences in outcomes are unlikely to be driven by gross diagnostic imbalance alone.

Using complementary analytic approaches, risk-standardized models and observed-to-expected (O/E) ratios—we found no meaningful regional differences in adjusted mortality, reinforcing the notion that surgical outcomes for ACHD patients are broadly comparable nationwide once patient complexity is accounted for. However, resource utilization exhibited consistent regional variation. The Northeast repeatedly demonstrated the lowest adjusted and observed hospitalization costs, with narrow confidence intervals, suggesting more predictable and efficient resource use. In contrast, the Midwest and South showed higher-than-expected HRU rates, highlighting potential inefficiencies that are not fully explained by case mix alone.

Importantly, while lengths of stay were largely comparable across regions, cost differences persisted even after adjustment, indicating that factors beyond duration of hospitalization—such as intensity of care, ancillary services, and institutional practice patterns—may play a substantial role.

Procedure-specific analyses reinforced these findings. Heart transplantation was associated with the highest mortality and resource use across all regions, as expected, but without statistically significant regional differences in mortality. For valve repair, valve replacement, and CABG, mortality remained low and comparable across regions, yet costs and HRU varied substantially, particularly with higher utilization in the South and lower costs in the Northeast. These findings suggest that regional practice patterns and system-level factors contribute to residual variation beyond procedural complexity alone.

Consistent with prior literature, patient complexity remains a key driver of resource utilization. For instance, Benderly et al. reported the highest standardized service utilization ratios among patients with complex congenital heart disease.(10) Similarly, Bhatt et al. found that higher surgical complexity doubled the likelihood of HRU admissions, defined as exceeding the 90th percentile for hospital charges.(11) In our adjusted model, clinical complexity—captured by conotruncal anatomy, comorbidity burden (Elixhauser index), and operative risk (RACHS-2)—was independently associated with higher hospitalization costs. Although granular clinical data were unavailable, these validated proxy measures provided robust adjustment for comorbidity and surgical risk. (12,13) Likewise, excluding congenital aortic insufficiency—a lower-risk subgroup—resulted in uniformly higher mortality, HRU, and cost estimates.

Beyond patient complexity, institutional characteristics and market dynamics likely contribute to observed variation. (14,15) The expansion of Adult Congenital Heart Association (ACHA)-accredited programs, from 5 in 2016 to approximately 40 by 2020,(16) may have influenced regional cost profiles by altering referral concentration, care pathways, and competitive pressures. Regions with higher density of accredited centers, such as the Northeast, may benefit from greater specialization and operational efficiency.(17)

Additionally, regional insurance market concentration may indirectly shape hospital behavior. In highly concentrated insurance markets, lower reimbursement rates may incentivize hospitals to adopt cost-containment strategies that influence observed cost patterns. (18,19) While negotiated prices are distinct from hospital costs, these market forces can affect staffing models, resource allocation, and procedural decision-making in ways not directly captured by administrative data.

This study represents a contemporary national evaluation of regional variation in resource utilization and costs among ACHD patients undergoing cardiac surgery. Importantly, it demonstrates that equivalent survival outcomes coexist with substantial differences in resource utilization between regions, shifting the focus of quality improvement from mortality alone toward value-based care metrics. Following the framework proposed by Kilbourne et al., this work constitutes the foundational “detection” phase of disparities research, systematically identifying where variation exists. (20) Future studies should move toward the “understanding” phase by disentangling the contributions of institutional practices, care pathways, referral networks, and market forces—and ultimately toward the “intervention” phase by designing targeted strategies to optimize care delivery in higher-utilization regions.

While comprehensive, this study has limitations. The HCUP database relies on ICD-10 codes, which may not fully capture procedural complexity, and lacks granular data on clinical severity. This restricts the ability to assess the physiological burden of congenital cardiopathies or the cumulative impact of multiple interventions over time. The absence of more granular data affects the robustness of risk adjustment and our ability to fully capture the interactions between regional variation and outcomes. Future studies incorporating clinical registries with detailed anatomical and procedural data are needed to refine these findings and inform more targeted policies.

Moreover, without access to longitudinal data—such as morbidity and mortality, and outpatient care burden—we are unable to fully evaluate the efficacy of care or identify areas where resource utilization may not translate to improved patient outcomes and comprehensively assessing of value in ACHD care.

## Conclusion

In this national cohort of ACHD patients undergoing cardiac surgery, regional differences in mortality were minimal after adjustment, but substantial variation persisted in hospitalization costs and resource utilization. These findings suggest that unmeasured factors— including both system-level and patient complexity—drive much of the observed regional variation. As the U.S. healthcare system moves toward value-based care, recognizing these regional and complexity-driven patterns will be essential for designing targeted strategies that balance cost containment with equitable access and high-quality care for this growing and resource-intensive population.

## Abbreviations

ACHD: adults with congenital heart disease
CABG: coronary artery bypass graft
CHD: congenital heart disease
ECMO: extracorporeal membrane oxygenation
GLM: generalized linear model
HCUP: Healthcare Cost and Utilization Project
HRU: High resource utilization
ICD-10-CM: International Classification of Disease, 10^th^ revision, Clinical Modification
IQR: interquartile range
LOS: length of stay
US: United States

## Data Availability

Data will be available upon reasonable request.

